# Overestimation of anticoagulant benefit in patients with atrial fibrillation and low life expectancy: evidence from 12 randomized trials

**DOI:** 10.1101/2023.02.10.23285303

**Authors:** Sachin J. Shah, Carl van Walraven, Sun Young Jeon, W. John Boscardin, FD Richard Hobbs, Stuart Connolly, Michael Ezekowitz, Kenneth E. Covinsky, Margaret C. Fang, Daniel E. Singer

## Abstract

**Background:** Patients with atrial fibrillation (AF) have a high rate of all-cause mortality that is only partially attributable to vascular outcomes. While the competing risk of death may affect expected anticoagulant benefit, guidelines do not account for it. We sought to determine if using a competing risks framework materially affects the guideline-endorsed estimate of absolute risk reduction attributable to anticoagulants.

**Methods:** We conducted a secondary analysis of 12 RCTs that randomized patients with AF to oral anticoagulants or either placebo or antiplatelets. For each participant, we estimated the absolute risk reduction (ARR) of anticoagulants to prevent stroke or systemic embolism using two methods. First, we estimated the ARR using a guideline-endorsed model (CHA_2_DS_2_-VASc) and then again using a Competing Risk Model that uses the same inputs as CHA_2_DS_2_-VASc but accounts for the competing risk of death and allows for non-linear growth in benefit over time. We compared the absolute and relative differences in estimated benefit and whether the differences in estimated benefit varied by life expectancy.

**Results:** 7933 participants had a median life expectancy of 8 years (IQR 6, 12), determined by comorbidity-adjusted life tables. 43% were randomized to oral anticoagulation (median age 73 years, 36% women). The guideline-endorsed CHA_2_DS_2_-VASc model estimated a larger ARR than the Competing Risk Model (median ARR at 3 years, 6.9% vs. 5.2%). ARR differences varied by life expectancies: for those with life expectancies in the highest decile, 3-year ARR difference (CHA_2_DS_2_-VASc model – Competing Risk Model 3-year risk) was −1.2% (42% relative underestimation); for those with life expectancies in the lowest decile, 3-year ARR difference was 5.9% (91% relative overestimation).

**Conclusion:** Anticoagulants were exceptionally effective at reduced stroke risk. However, anticoagulant benefits were misestimated with CHA_2_DS_2_-VASc, which does not account for the competing risk of death nor decelerating treatment benefit over time. Overestimation was most pronounced in patients with the lowest life expectancy and when benefit was estimated over a multi-year horizon.

## INTRODUCTION

Anticoagulants are the mainstay of preventative therapy for millions of older adults with atrial fibrillation. While anticoagulants reduce the risk of ischemic stroke and systemic embolism, they also increase the risk of bleeding. To help patients and clinicians weigh the risks and benefits of treatment, clinical guidelines and decision support tools endorse using the CHA_2_DS_2_-VASc score to estimate a patient’s annual risk of ischemic stroke or systemic embolism without treatment.^1–5^ The ACC/AHA/HRS consensus guidelines recommend that this baseline risk be used to estimate a patient’s expected absolute risk reduction by applying the relative risk reduction from a meta-analysis of randomized trials.^1,6^ Guidelines recommend anticoagulant therapy when the absolute risk reduction exceeds a threshold. The premise is that treatment benefits outweigh risks when an individual’s estimated event risk exceeds this threshold.

While transparent, this approach makes two assumptions that can affect the accuracy of expected benefit. First, this approach does not account for the competing risk of death. A competing risk is an alternative outcome that occurs before, and necessarily precludes, the event of interest (e.g., cancer death before stroke from atrial fibrillation), thus limiting the absolute benefit achievable by anticoagulant treatment.^7–11^ Competing risks are germane given the high all-cause mortality rate following a new diagnosis of atrial fibrillation—multiple studies estimate 20-25% mortality in the first year.^12–14^ Second, this approach assumes that the therapeutic benefit continues to increase at a constant rate over time—that is, the risk of stroke over two years is twice the one-year risk and thus the absolute risk reduction over two years is twice the one-year absolute risk reduction. Both issues are readily addressed by estimating benefit using a competing risk model; however, it is unknown if doing so will materially affect absolute risk reduction estimates attributable to anticoagulants.

We used patient-level data from 12 randomized trials of anticoagulants for atrial fibrillation to determine if a competing risk model affects the measurement of absolute stroke risk reduction. First, we compared the guideline-endorsed approach to measuring absolute risk reduction (i.e., CHA_2_DS_2_-VASc score) to a competing risk model (i.e., Fine-Gray model^15^). Second, we determined if differences in expected stroke risk reduction between the guideline-endorse CHA_2_DS_2_-VASc approach and a competing risk model varied by life expectancy.

## METHODS

### Study design and participants

We used patient-level data from the Atrial Fibrillation Investigators (AFI) database which contains patient-level data from 12 published clinical trials where patients were randomized to full-dose oral anticoagulant, antiplatelets, or placebo. We focused on trials that established the efficacy of oral anticoagulants; therefore, we did not include trials that compared two different oral anticoagulants. We included the following trials: Atrial Fibrillation, Aspirin, and Anticoagulation Study 1 (AFASAK-1),^16^ AFASAK-2,^17^ Boston Area Anticoagulation Trial for Atrial Fibrillation (BAATAF),^18^ Birmingham Atrial Fibrillation Treatment of the Aged Study (BAFTA),^19^ Canadian Atrial Fibrillation Anticoagulation (CAFA),^20^ European Atrial Fibrillation Trial (EAFT),^21^ Primary Prevention of Arterial Thromboembolism in Atrial Fibrillation (PAATAF),^22^ National Study for Prevention of Embolism in Atrial Fibrillation (NASPEAF),^23^ the Stroke Prevention in Atrial Fibrillation 1 (SPAF-1),^24^ SPAF-2,^25^ SPAF-3,^26^ and Stroke Prevention in Non-rheumatic Atrial Fibrillation (SPINAF).^27^ We did not include patients with mitral stenosis and patients in SPAF-1, EAFT, PATAF, and SPAF-3 who were deemed ineligible to receive anticoagulants (trial details in **Appendix 1** and **Appendix 2**).

### Participant characteristics

Research coordinators and physicians collected patient characteristics before therapy initiation. While specific features varied from study to study, common elements included a history of stroke or transient ischemic attack, hypertension or systolic blood pressure ≥160 mmHg or use of antihypertensives, diabetes, angina, myocardial infarction, peripheral vascular disease, smoking, and congestive heart failure, and body mass index. History of myocardial infarction was not collected in NASPEAF and peripheral vascular disease was not collected in AFASAK1, BAATAF, or BAFTA. Because history of myocardial infarction and peripheral vascular disease were missing for all participants in specific trials, we assumed they were missing at random and imputed them in 20 datasets using chained equations.^28^ We excluded <1% of participants who were missing data otherwise collected in a given trial (**Appendix 3**).

### Treatment exposure

We examined all patients based on their treatment allocation (i.e., intention to treat). Because studies have shown that antiplatelet and low-dose warfarin are ineffective thromboprophylaxis in AF,^6^ we categorized all trial participants as being randomized to full-dose anticoagulants or control. Patients randomized to placebo, antiplatelets, low-dose warfarin, or low-dose warfarin with aspirin were considered controls. AFASAK2, PAATAF, and SPAF3 used low-dose warfarin and reported a mean international normalized ratio (INR) of < 1.5 supporting their categorization as a control. While NASPEAF also had a low-dose anticoagulant arm, the mean INR was 2.0 which is considered therapeutic^1^; therefore, we excluded participants randomized to the NASPEAF low-dose anticoagulant arm.

### Outcome ascertainment

The primary outcome was ischemic stroke or systemic embolism. We detail outcome definitions by trial in **Appendix 4**. In general, trials defined ischemic stroke as a focal neurological deficit lasting >24 hours. All trials except AFASAK-1 required a CT or MRI showing the absence of blood. Systemic embolism was collected as an outcome in all but SPINAF and, by and large, defined as an embolism to internal organs or limbs and required evidence via angiography, surgery, or autopsy (**Appendix 4**). Patients were evaluated at 3- to 6-month intervals or when a clinical outcome event was suspected. Except in AFASAK-1, a central committee, blinded to intervention allocation, adjudicated all clinical events.

### Life expectancy

We estimated the life expectancy of each participant at the time of trial enrollment using the life table method.^29^ We started with gender- and enrollment year-specific life tables from the U.S. Centers for Disease Control and Prevention.^30^ These tables, generated from population data, predict annual mortality rates stratified by age, sex, and year. The life table method uses annual mortality rates to calculate life expectancy. We used the life table method to estimate each participant’s life expectancy by adjusting the annual mortality rate for the additional mortality risk associated with their comorbidities at the time of trial enrollment (**Appendix 5**).

### Analysis

Our first analytic goal was to determine if using a competing risk framework generates estimates of stroke risk reduction different from those of the guideline-endorsed CHA_2_DS_2_-VASc model. To accomplish this and speak directly to guideline-recommended practice, we estimated absolute risk reduction using the CHA_2_DS_2_-VASc score as the ACC/AHA/HRS guidelines recommend.^1,2^ Specifically, we assigned each patient an off-treatment risk of ischemic stroke or systemic embolism corresponding to their CHA_2_DS_2_-VASc score. Rates come from the 2012 study by Friberg et al., which used the Swedish Atrial Fibrillation cohort to validate off-treatment thromboembolic rates corresponding to each CHA_2_DS_2_-VASc score.^31^ These rates are used in patient-facing decision tools, online calculators, and decision analytic models.^5,32,33^ To calculate the absolute risk reduction, we multiplied the off-treatment stroke rate by 0.64, the guideline-cited efficacy of anticoagulants.^1,6^ Using this procedure for each patient, we estimated the annual absolute risk reduction; this precise method is endorsed by the ACC/AHA/HRS Atrial Fibrillation management guidelines to estimate benefit.^1^ Because patients and physicians prefer to make anticoagulant decisions using a 1-to-5-year time horizon,^5^ we extrapolated this annual reduction over five years, accounting for the declining at-risk population (**Appendix 6**).^34,35^ The same approach is also used in decision aids.^5^

Next, we estimated the absolute risk reduction using the Fine-Gray extension of the Cox proportional hazards model, treating death unrelated to ischemic stroke or systemic embolism as a competing event.^11^ We fit a Fine-Gray model where time to ischemic stroke or systemic embolism is a function of age (<65 years, 65-74 years, >75years), gender, congestive heart failure, diabetes, hypertension, prior stroke or transient ischemic attack, and vascular disease stratified by randomization to oral anticoagulants. These the same predictors used in the CHA_2_DS_2_-VASc score. Then, we used the resulting treatment-stratified models to estimate the cumulative incidence of ischemic stroke or systemic embolism for each participant given their covariates at each study time point, assuming first they had been randomized to oral anticoagulants and then assuming they had been randomized to control (i.e., predicted values).^36,37^ The difference between the two estimates represented the ARR for a given patient at a given time point. We determined the misestimation of the CHA_2_DS_2_-VASc method as the difference between the ARR estimated by CHA_2_DS_2_-VASc and the ARR estimated by the Competing Risk Model. We used the paired t-test to determine if the two methods produced statistically different estimates of benefit at each year after randomization.

Our second analytic goal was to determine if life expectancy predicted magnitude in the CHA_2_DS_2_-VASc model misestimation. To achieve this, we determined the association between life expectancy and misestimation of the CHA_2_DS_2_-VASc method over a 3-year horizon. We chose 3 years since it is the midpoint between the 1-to-5 year horizon preferred by patients and physicians and has been used in prior anticoagulation decision analyses.^5,38^ We examined misestimation of the CHA_2_DS_2_-VASc method by decile of life expectancy at trial enrollment, hypothesizing that the misestimation would be greater at lower life expectancies.

We performed all statistical analyses using SAS 9.4 (Cary, NC). The study protocol was approved by Institutional Review Boards at UCSF (21-34930) and MGH (2022P001783).

## RESULTS

### Patient characteristics and overall event rates

This study included 7933 patients from 12 randomized trials where 3407 (43%) were randomized to oral anticoagulation (**Table 1**). The median age was 73 years at enrollment, 36% were women, and the median CHA_2_DS_2_-VASc score was 3 [IQR 2, 4]. At enrollment, the median life expectancy was 8 years [IQR 6, 12] (**Appendix 7**). Most patients (83%) ended the follow-up period without a clinical event (median 731 days; IQR 415, 1025). (**Table 2**). In these trials, 530 (7%) patients’ first clinical event was an ischemic stroke or systemic embolism (median 334 days; IQR 120, 580). Additionally, 630 (8%) patients died before a stroke or systemic embolism (median 457 days; IQR 216, 772).

**Table 1:** Characteristics of patients with atrial fibrillation in 12 randomized trials. AFASAK - Atrial Fibrillation, Aspirin, and Anticoagulation Study; BAATAF - Boston Area Anticoagulation Trial for Atrial Fibrillation; BAFTA - Birmingham Atrial Fibrillation Treatment of the Aged Study; CAFA - Canadian Atrial Fibrillation Anticoagulation; EAFT - European Atrial Fibrillation Trial; PAATAF - Primary Prevention of Arterial Thromboembolism in Atrial Fibrillation; NASPEAF - National Study for Prevention of Embolism in Atrial Fibrillation; SPAF - Stroke Prevention in Atrial Fibrillation; SPINAF - Stroke Prevention in Non-rheumatic Atrial Fibrillation * History of myocardial infarction was not available for NSPEAF trial participants † History of peripheral vascular disease was not available for AFASAK1, BAATAF, or BAFTA trial participants ‡ Height and weight were not available for BAATAF or AFASAK2 trial participants § Smoking status was not available for AFASAK1, CAFA, or BAFTA participants. In EAFT, data collection did not distinguish between former and never smokers. CHA_2_DS_2_-VASc scores are for those with complete cases. It excludes 2395 participants enrolled in trials where peripheral vascular disease data are not available, and 543 participants enrolled in trials where history of myocardial infarction was not available. ¶ Control includes those assigned placebo, aspirin, low-dose warfarin, or low-dose warfarin an aspirin. Patients enrolled in SPAF3 and AFASAK2 and assigned to low-dose warfarin had a mean internal normalized ratio (INR) of < 1.5 supporting their categorization as a control. Patients enrolled in NASPEAF and randomized to low-dose anticoagulant had a mean INR of 2.0 and were therefore not included as a control. Since missing data were missing for entire trials, they were assumed to be missing completely at random.

| **Characteristic** | **n=7933** |
| --- | --- |
| Age, years (median [IQR]) | 73 [67, 78] |
| Gender |  |
| Male | 5040 (64%) |
| Female | 2893 (36%) |
| Diabetes |  |
| No | 6780 (85%) |
| Yes | 1153 (15%) |
| Hypertension |  |
| No | 4049 (51%) |
| Yes | 3884 (49%) |
| Congestive heart failure |  |
| No | 5533 (70%) |
| Yes | 2400 (30%) |
| Prior stroke |  |
| No | 6413 (81%) |
| Yes | 1520 (19%) |
| Angina |  |
| No | 6566 (83%) |
| Yes | 1367 (17%) |
| Prior myocardial infarction |  |
| No | 6514 (82%) |
| Yes | 876 (11%) |
| Missing* | 543 (7%) |
| Peripheral vascular disease |  |
| No | 5109 (64%) |
| Yes | 429 (5%) |
| Missing<sup>†</sup> | 2395 (30%) |
| Body mass index kg/m<sup>2</sup> (median [IQR]) |  |
| BMI among those with data | 26 [24, 29] |
| Missing<sup>‡</sup> | 2070 (26%) |
| Smoking status |  |
| Never smoker | 2427 (31%) |
| Former smoker | 1843 (23%) |
| Current smoker | 790 (10%) |
| Missing<sup>§</sup> | 2873 (36%) |
| CHA<sub>2</sub>DS<sub>2</sub>-VASc score |  |
| Score (median [IQR]) | 3 [2, 4] |
| Missing<sup>-</sup> | 2938 (37%) |
| Trial |  |
| AFASAK1 | 1002 (13%) |
| AFASAK2 | 677 (9%) |
| BAATAF | 420 (5%) |
| BAFTA | 973 (12%) |
| CAFA | 375 (5%) |
| EAFT | 661 (8%) |
| NASPEAF | 543 (7%) |
| PATAF | 364 (5%) |
| SPAF1 | 208 (3%) |
| SPAF2 | 1098 (14%) |
| SPAF3 | 1044 (13%) |
| SPINAF | 568 (7%) |
| Randomized study arm assignment |  |
| Warfarin | 3407 (43%) |
| Control<sup>¶</sup> | 4526 (57%) |

**Table 2:** Trial outcomes, rates and follow-up time.

| **First clinical outcome** | **Events (%)** | **Days from randomization,**<br>**median (IQR)** |
| --- | --- | --- |
| Ischemic stroke or systemic embolism | 530 (7%) | 334 (120, 580) |
| Systemic bleed | 175 (2%) | 413 (163, 731) |
| Intracranial hemorrhage | 29 (0%) | 300 (201, 620) |
| Death, all-cause | 630 (8%) | 457 (216, 772) |
| Study end without a clinical event | 6569 (83%) | 731 (415, 1025) |

### Comparison of absolute risk reduction estimates

Relative to the Competing Risk Model, the CHA_2_DS_2_-VASc model overestimated the absolute risk reduction (ARR) of anticoagulants (**Figure 1**). As the time horizon increased, the CHA_2_DS_2_-VASc estimate of median benefit increased linearly. In contrast, the Competing Risk Model estimated a non-linear absolute risk reduction over time—while benefit increased over time, it decelerated, i.e., absolute risk reduction grew by less each year. As a result, the CHA_2_DS_2_-VASc model overestimated anticoagulation benefit as the time-horizon increased. After 1 year, the CHA_2_DS_2_-VASc model and the Competing Risk Model produced clinically similar estimates of absolute risk reduction (median ARR 2.3% by CHA_2_DS_2_-VASc estimate vs. 2.4% by Competing Risk Model, p<0.001). After 3 years, the CHA_2_DS_2_-VASc-based ARR was clinically and statistically larger than the ARR from the Competing Risk Model (median ARR 6.9% vs. 5.2%, p<0.001). This difference increased when absolute risk reduction was estimated over a 5-year horizon (median ARR 11.2% vs. 6.3%, p<0.001).

**Figure 1:**
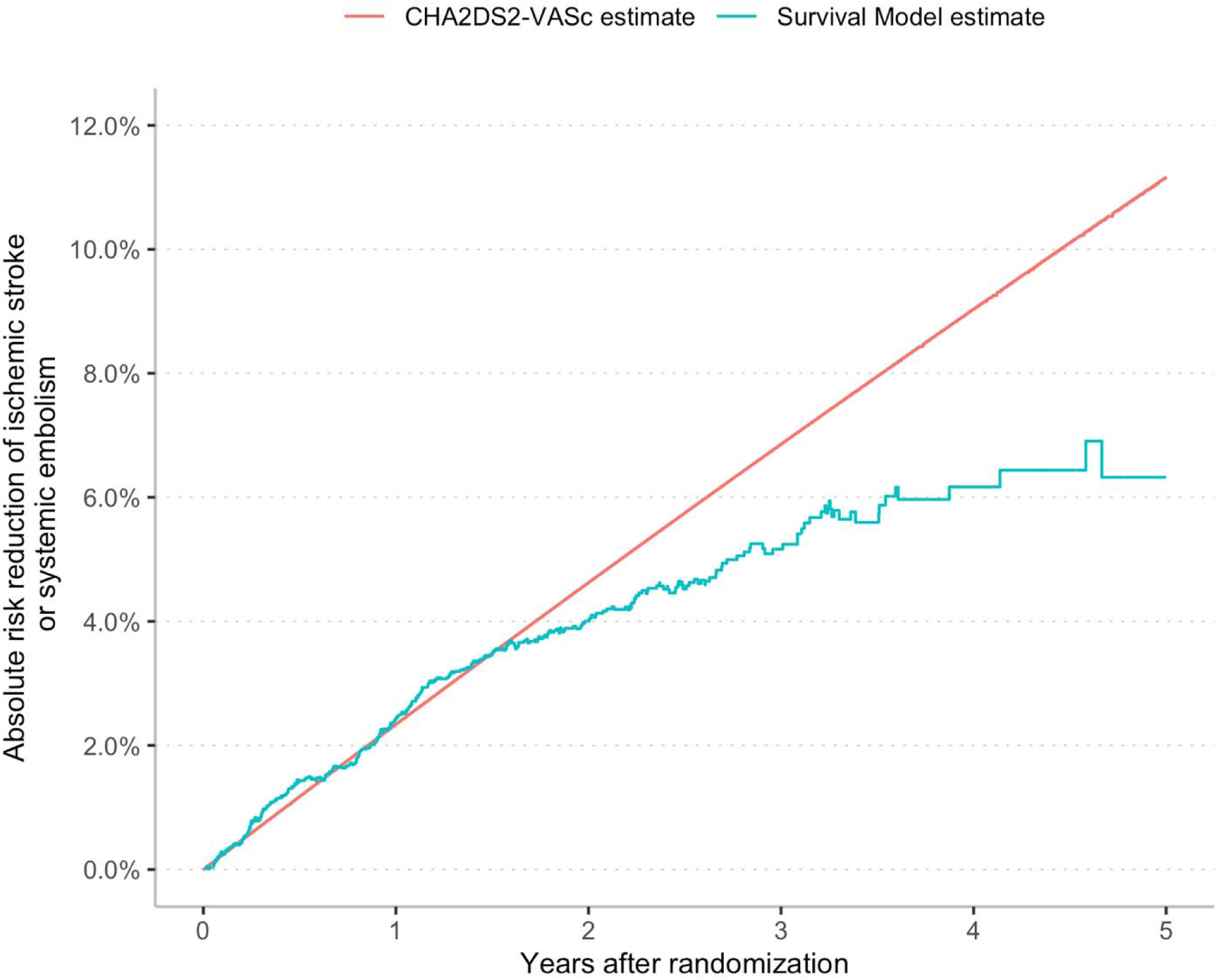
Estimated median absolute risk reduction by CHA_2_DS_2_-VASc model compared with Competing Risk Model. For each patient in the cohort, we estimate the absolute risk reduction attributable to oral anticoagulation annually for 5 years using the CHA_2_DS_2_-VASc model and again using a Fine-Gray model, a survival model that accounts for the competing risk of death. We plot the median benefit at each time point. We graphed the median ARR because the CHA_2_DS_2_-VASc model produces discrete estimates of benefit (i.e., not normally distributed). Data are presented as a table in **Appendix 10**. Component on- and off-treatment cumulative incidence rates also displayed in **Appendix 10**.

We assess whether the observed discrepancy in ARR could be because the guidelines estimate of off-treatment stroke risk were produced in an external cohort and therefore were miscalibrated (**Appendix 9**). The sensitivity showed that recalibration to the AFI database did not meaningfully change the results presented in **Figure 1**.

### Life expectancy and misestimation of benefit

As life expectancy decreased, the CHA_2_DS_2_-VASc model increasingly overestimated the stroke and systemic embolism risk reduction attributable to anticoagulants in absolute and relative terms (**Figure 2**). The figure plots the absolute and relative difference between the 3-year ARR estimated by the CHA_2_DS_2_-VASc model and the Competing Risk Model by life expectancy decile at trial enrollment. In the decile with the highest life expectancy (16 to 47 years), on average, over 3 years, the CHA_2_DS_2_-VASc model underestimated benefit by 1.2% (95% CI 1.1% to 1.3%) in absolute terms and 42% (95% CI 40% to 45%) in relative terms. By comparison, in the decile with the lowest life expectancy (1 to 4 years), on average over 3 years, the CHA_2_DS_2_-VASc model overestimated benefit by 5.9% (95% CI 5.6% to 6.1%) in absolute terms and 91% (95% CI 87% to 95%) in relative terms.

**Figure 2:**
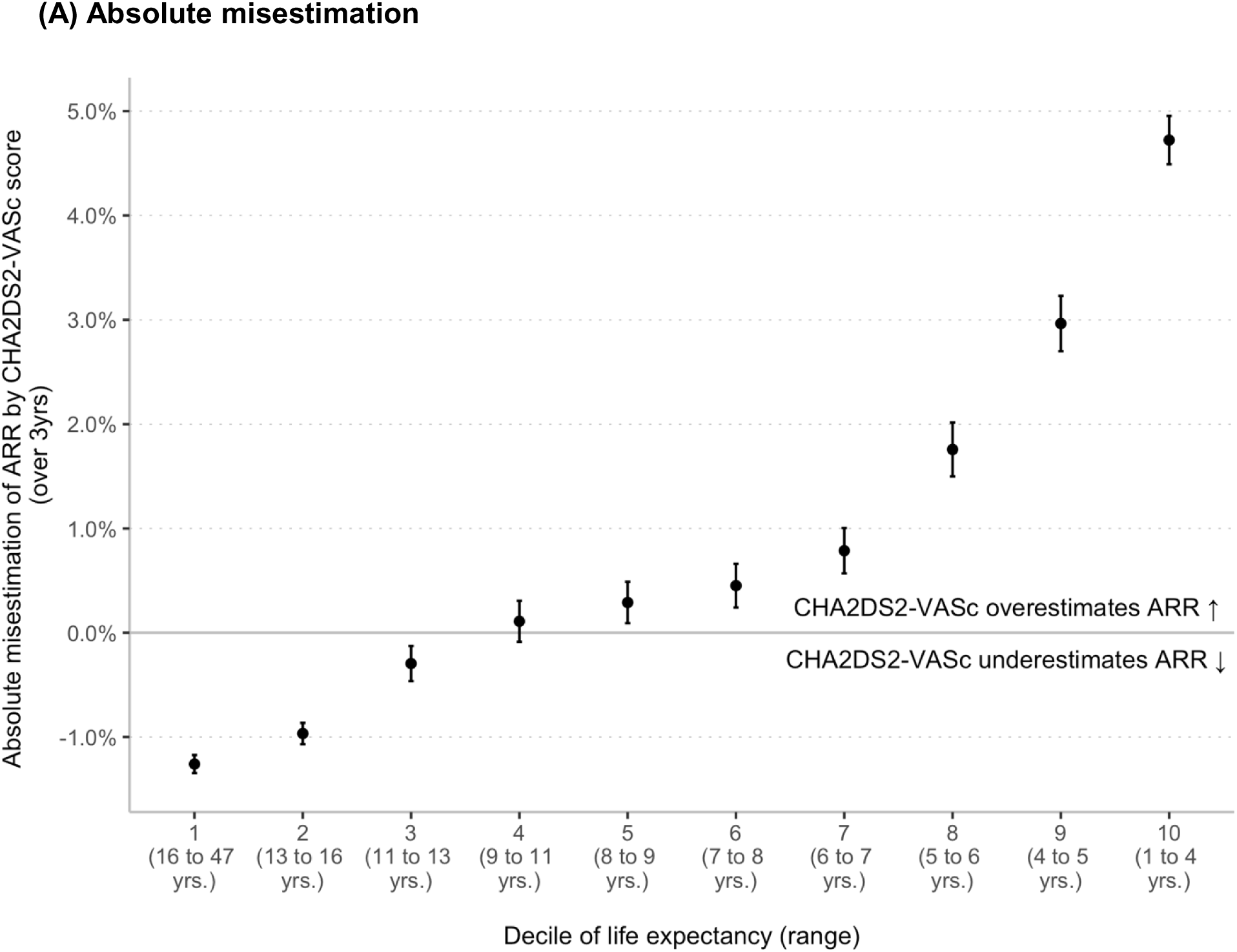
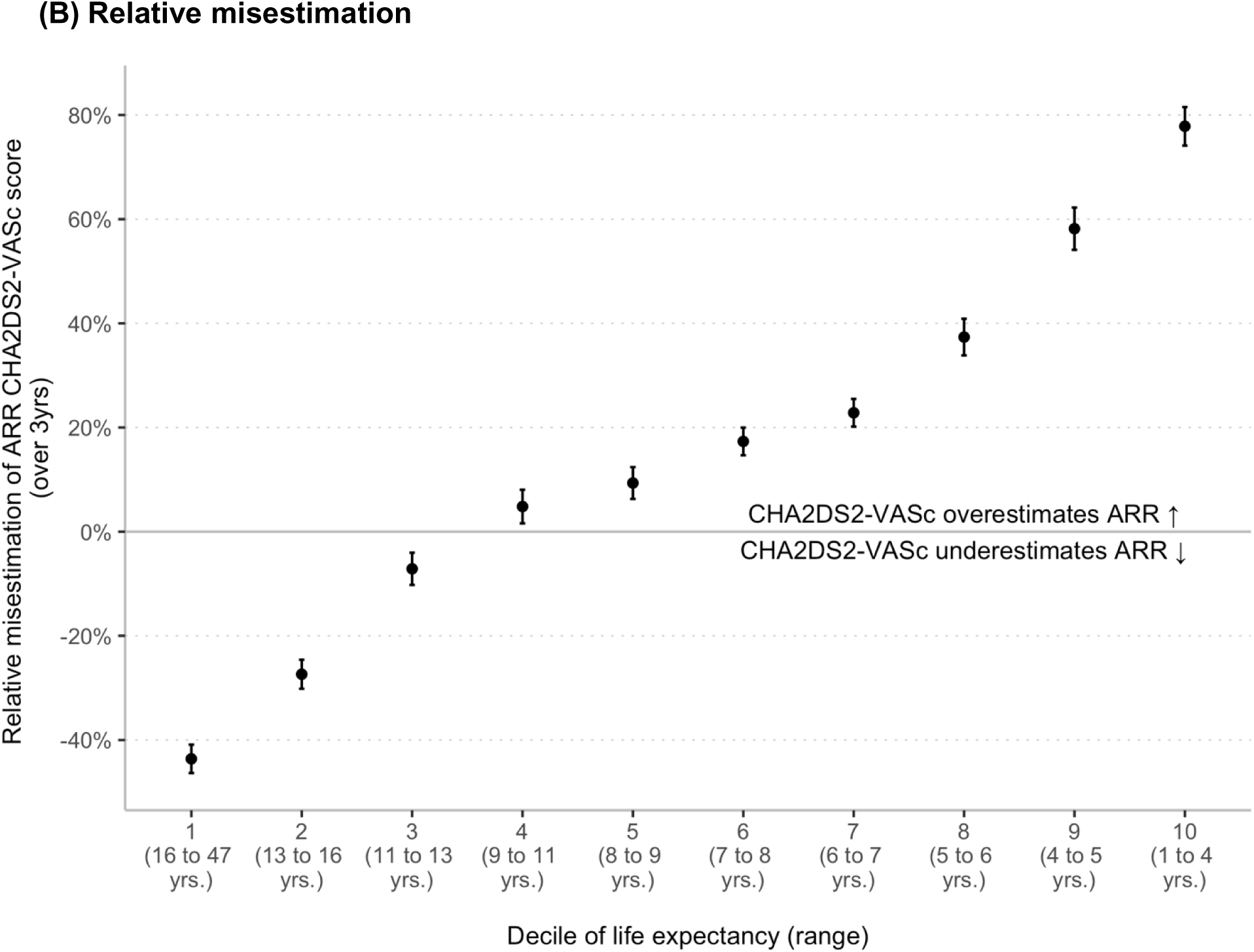
Misestimation of Stroke Risk Reduction by CHA_2_DS_2_-VASc score at 3 years, Absolute misestimation is defined as: ARR_CHA2DS2-VASc_ – ARR_Competing Risk Model_. Relative misestimation is defined as: ARR_CHA2DS2-VASc_ / ARR_Competing Risk Model_ – 1. The dot represents the mean overestimation, and the error bars represent the 95% confidence interval of the mean. The misestimation of absolute risk reduction (ARR) is calculated for each patient as the difference between the ARR computed by the CHA_2_DS_2_-VASc score and the ARR computed by the Fine-Gray model, a survival model that accounts for the competing risk of death. Positive numbers represent the overestimation of the CHA_2_DS_2_-VASc model. Tabular results can be found in **Appendix 11.**

## DISCUSSION

Using patient-level data from 12 randomized trials, we demonstrated that while anticoagulants effectively reduce ischemic stroke and systemic embolism risk, failing to use a competing risks framework resulted in a meaningful overestimation of treatment benefit. This finding was most pronounced when risk reduction was estimated over a multi-year horizon. Further, we showed that as life expectancy decreased, treatment benefit was increasingly overestimated. While those with the highest life expectancy may benefit more than guideline estimates would suggest, benefit for those with the lowest life expectancy was strikingly overestimated.

The study results directly apply to the AHA/ACC/HRS^1,2^ and European Society of Cardiology (ESC)^3^ atrial fibrillation guidelines in which the cornerstone of anticoagulant decision-making is estimating the absolute risk reduction. Guidelines ask clinicians to estimate the off-treatment stroke risk using the CHA_2_DS_2_-VASc score and to use that baseline risk to infer the probable absolute risk reduction. Anticoagulants are recommended above a CHA_2_DS_2_-VASc score threshold—i.e., when the absolute risk reduction exceeds a threshold. Thus, if clinical guidelines continue to recommend treatment using an absolute risk reduction threshold, these results suggest guidelines should re-estimate benefit using a competing risk framework. At the very least, guidelines should acknowledge that current methods overestimate benefits for those with limited life expectancy and when estimating benefits over a multi-year horizon.

The current study’s findings should influence anticoagulant decision aids for patients with atrial fibrillation. To advance anticoagulant shared decision-making, investigators and professional societies have developed conversation aids that display a patient’s risk of stroke with and without anticoagulants. For example, the American College of Cardiology’s CardioSmart tool and the Mayo Clinic Anticoagulation Choice Decision Aid both display a pictogram of absolute risk with and without treatment to communicate treatment effects.^4,5^ These implementation tools are built to reflect clinical guidelines and do so faithfully. However, both should note that benefit estimates are overstated for those with limited life expectancy and when benefit is estimated over a multi-year horizon.

These results also lend credence to physicians for whom advanced age, frailty, and function—all significant determinants of life expectancy—factor in their anticoagulant decision-making.^39,40^ Adults aged 65 years and older constitute 80% of all American adults with atrial fibrillation.^41^ Further, prior work indicates that many older adults with atrial fibrillation have geriatric syndromes (e.g., dependency in activities of daily living) known to be associated with a reduced life expectancy.^42,43^ In this study, we estimated life expectancy using basic medical comorbidity data available in the trial database. Modern, more accurate tools go beyond comorbidities using physical function, cognition, and self-reported health to estimate life expectancy.^44,45^ Life expectancy estimates from such tools are routinely used to inform the risk and benefits of interventions in older adults (e.g., cancer screening). Until guidelines formally account for it, clinicians may consider using life expectancy to guide the discussion about the benefit of anticoagulants when treating older adults. This may be particularly relevant when treating patients with both a limited life expectancy and borderline CHA_2_DS_2_-VASc scores.

Finally, these findings should inform the methods used to develop and validate future stroke risk models. While the CHA_2_DS_2_-VASc score continues to be endorsed by U.S. and European guidelines, investigators are actively developing a new generation of stroke risk prediction models. Contemporary models like the ABC stroke risk score, CARS, and the ATRIA stroke model all outperform the CHA_2_DS_2_-VASc score.^46–48^ However, none used an analysis framework that both accounts for the competing risk of death and changing risk over a multi-year horizon. The ABC model establishes a non-linear stroke risk by showing 3-year risk is not simply three times the 1-year risk, findings that were redemonstrated in this paper. When developing the ABC model, Hijazi et al. also conducted a sensitivity analysis comparing their model to a competing risk model.^46^ They found a tight correlation when risk was estimated over a 1-year horizon, results mirrored in this study. We expanded on their work by showing that this correlation was weaker when using a longer time horizon (e.g., 3 years). More importantly, we identified substantial heterogeneity—as life expectancy decreased, overestimation increased.

There are limitations to this study inherent to the data available and the study design. First, this study relied on data from RCTs conducted between 1989 and 2007 and thus may be only partially representative of contemporary patients with atrial fibrillation. Specific differences include the risk of stroke and death from non-atrial fibrillation causes and the added safety of direct-acting anticoagulants.^49^ This limitation is balanced by the fact that the AFI cohort is one of the largest patient-level atrial fibrillation cohorts where anticoagulant treatment was randomized against placebo and antiplatelets. The results were unaffected by selection bias that hampers contemporary risk models developed in observational cohorts. More importantly, while dated, these trials are the foundation upon which current guidelines recommend anticoagulants. Second, this study could not address the relationship between life expectancy and the potential misestimation of anticoagulant harm. Specifically, the AF Investigators database does not include inputs used in contemporary hemorrhage prediction tools (e.g., ATRIA bleed, HAS-BLED). While it is important to consider the effect a competing risk framework may have on estimating the risk of bleeding, current guidelines do not incorporate bleeding risk into treatment recommendations. For example, for patients with scores above the CHA_2_DS_2_-VASc treatment threshold, guideline recommendations do not change whether the bleeding risk is high or low. Finally, because nationality was unavailable for study participants, we relied on U.S. life tables to calculate life expectancy.

In summary, we showed that while oral anticoagulants were clearly effective, treatment benefit was overstated when using the guideline-endorsed approach because guidelines do not account for the competing risk of death and assume a constant growth in treatment benefit over time. Overestimation was most pronounced in patients with the lowest life expectancy and when benefit was estimated over a multi-year horizon. These findings should inform guidelines and decision aids, clinicians treating patients with limited life expectancy, and investigators developing stroke risk prediction models.

## Data Availability

Researchers can apply to the AF Investigators for data access.

## ACKNOWLEDGMENTS

We thank Dr. Sei Lee, Professor of Medicine at UCSF, for his valuable methodological feedback. Dr. Shah had full access to all the data in the study and takes responsibility for the integrity of the data and the accuracy of the data analysis.

## Author contributions

Dr. Shah had full access to all of the data in the study and takes responsibility for the integrity of the data and the accuracy of the data analysis. All authors listed have contributed sufficiently to the project to be included as authors, and all those who are qualified to be authors are listed in the author byline.

## Data sharing

Researchers can apply to the AF Investigators for data access.

