## appendix for "Overestimation of anticoagulant benefit in patients with atrial fibrillation and low life expectancy: evidence from 12 randomized trials"

### Appendix 1: Trial inclusion and exclusion criteria and study period

| **Trial** | **Inclusion** | **Exclusion** | **Trial period** |
| --- | --- | --- | --- |
| Atrial Fibrillation, Aspirin, and Anticoagulation Study 1 (AFASAK-1)^15^ | ≥18 years old  ECG-verified atrial fibrillation  Primary physician consent | Anticoagulation for >6 months  Cerebrovascular event within the past month  Contraindications for aspirin or warfarin therapy  Previous side-effects of aspirin or warfarin therapy  Pregnancy or breastfeeding  Persistent BP > 180/100 mmHg  Psychiatric diseases  Chronic alcoholism  Heart surgery with valve replacement  Rheumatic heart disease | Nov 1985 to Jun 1988 |
| AFASAK-2^16^ | ≥18 years old  AF on 2+ ECGs at least 1 month apart | Age < 60 years with lone AF (i.e., with no underlying ischemic or hypertensive heart disease, congestive heart failure, hyperthyroidism, or chronic obstructive pulmonary disease)  Mitral stenosis  SBP >180 mm Hg  DBP >100 mm Hg  Stroke or TIA within 6 months  Risk factors for bleeding risk factors  Contraindication to warfarin or aspirin  Already receiving dose-adjusted warfarin  Mitral stenosis  Atrial flutter | May 1993 to Oct 1996 |
| Boston Area Anticoagulation Trial for Atrial Fibrillation (BAATAF)^17^ | AF on 2+ ECG  No evidence of mitral stenosis on echocardiography | Intermittent AF with no ECG with AF within 18 months  Transient AF during acute illness  Planned cardioversion  Echocardiographic evidence of intracardiac thrombus, ventricular aneurysm, severe congestive heart failure or prosthetic valve  Stroke <6 months ago predisposition to intracranial hemorrhage  TIA requiring treatment  Predisposition to intracranial hemorrhage  Indication or contraindication for anticoagulation  Required aspirin therapy  Abnormal thyroid function tests | Sept 1985 to Apr 1990 |
| Birmingham Atrial Fibrillation Treatment of the Aged Study (BAFTA)^18^ | Age ≥75 years; atrial fibrillation or atrial flutter on an ECG within the past 2 years | Rheumatic heart disease  Major nontraumatic hemorrhage within 5 years  Intracranial hemorrhage  Peptic ulcer disease by endoscopy in the last year  Esophageal varices  Allergy to warfarin or aspirin  Terminal illness as judged by primary care physician  Surgery within past 3 months  BP >180/110 mmHg  Primary care physician judged that the patient should or should not be on warfarin (i.e., no equipoise) | Apr 2001 to Sept 2006 |
| Canadian Atrial Fibrillation Anticoagulation (CAFA)^19^ | Chronic atrial fibrillation for ≥ 1 month or paroxysmal atrial fibrillation ≥3 times in previous 3 months (documented at least twice on ECG)  Age ≥19 years old  Absences of mitral valve prosthesis or mechanical aortic valve prosthesis  Absence of mitral valve stenosis on echocardiography | Requirement for anticoagulation  Contraindication to anticoagulation  Stroke or TIA within the past year  Requirement for antiplatelet drug therapy  Hyperthyroidism  Uncontrolled hypertension  Myocardial infarction within the past month | Jun 1987 to Apr 1990 |
| European Atrial Fibrillation Trial (EAFT)^20^ | Age >25 years old  TIA or minor stroke (grade 3 or less on modified Rankin scale) in previous 3 months  Atrial fibrillation at the time of TIA/stroke or paroxysmal atrial fibrillation in preceding 24 months  No rheumatic valvular disease on echocardiography | Atrial fibrillation secondary to other illness (e.g., hyperthyroidism)  Indication or contraindication for aspirin  NSAID medication use  Antiplatelet medication use  Oral anticoagulation use  No other cardioembolic source (e.g., prosthetic valve, cardiac aneurysm, atrial myxoma, cardiothoracic ration > 0.65, myocardial infarction in preceding 3 months, disorders of blood coagulation)  Scheduled for carotid endarterectomy or coronary surgery in next 3 months | Oct 1988 to Apr 1993 |
| Primary Prevention of Arterial Thromboembolism in Atrial Fibrillation (PAATAF)^21^ | Age ≥60 years  Chronic or intermittent atrial fibrillation on ECG in past 2 years | Treatable causes of AF  Previous stroke  Rheumatic valve disease  MI or cardiovascular surgery in past 1 year  Cardiomyopathy with left ventricular ejection fraction <40%  Chronic heart failure  Cardiac aneurysm  Previous systemic or retinal infarction  Coumarin use in past 3 months  Contraindications to aspirin or coumarin (hemoglobin concentration < 7.0 mmol/l, ventricular or duodenal ulcer in the past three years, gastrointestinal or urogenital bleeding in the past year, aspirin intolerance, coagulation disorder, and severe hepatic or renal disease)  Pacemaker  Life expectancy <2 years  Additional exclusion for standard anticoagulation: age ≥ 78 years, retinopathy, ventricular or duodenal ulcer, history of gastrointestinal hemorrhage, genitourinary bleeding, BP > 185/105 mm Hg | Jan 1990 to Dec 1996 |
| National Study for Prevention of Embolism in Atrial Fibrillation (NASPEAF)^22^ | Chronic or documented paroxysmal atrial fibrillation  Age > 60 years  High risk by SPAF3 criteria:   - Impaired left ventricle manifested by congestive heart failure within 100 days or fractional shortening ≤ 25% - Systolic blood pressure >160 mm Hg on one reading and >150 mm Hg on second reading or documented systolic blood pressure >160 mm Hg in preceding 3 months - Prior ischemic stroke, TIA, or systemic embolism more than 30 days prior to entry - Female and age over 75 years | Mechanical valve prosthesis  Stroke in previous 6 months  Serum creatinine >3 mg/dL  Alcoholism or drug addiction  Severe uncontrolled hypertension  Indication for NSAIDs  Indication or contraindication for antiplatelet or anticoagulant therapy | Jun 1995 to Jun 2001 |
| Stroke Prevention in Atrial Fibrillation 1 study (SPAF-1)^23^ | AF on ECG (constant or intermittent) in the preceding 12 months | Prosthetic heart valve  Personal physician refuses study  No echocardiographic evidence of rheumatic mitral stenosis  Successful cardioversion (electrical or chemical)  NYHA class IV congestive heart failure  Mitral regurgitation with congest heart failure and atrial diameter >5.5 cm  Idiopathic cardiomyopathy with congestive heart failure  Myocardial infarction in preceding 3 months  Coronary bypass surgery within previous 1 year  Coronary angioplasty within previous 3 months  Unstable angina within previous 1 year  Stroke, TIA, carotid endarterectomy in previous 24 months  Life expectancy < 24 months due to other medical condition  Chronic renal failure with serum creatinine > 3.0 mg/dL  Thrombocytopenia with platelet count < 100,000 platelets /mm^3^  Anemia with hemoglobin concentration < 10 g/dL  Severe chronic alcohol habituation  Indication or contraindication for warfarin or aspirin therapy  Treatment with non-steroidal anti-inflammatory drugs  Additional exclusion from anticoagulation arm:   - On antihypertensive therapy and SBP>180 mm Hg and DBP > 100 mm Hg - Prothrombin time > 2 seconds on 2 occasions - Stool test positive for occult blood - Inability to obtain follow up for prothrombin time monitoring - Previous intracranial hemorrhage, gastrointestinal, or genitourinary bleeding within previous 6 months - Previous severe hemorrhage with therapeutic anticoagulation - Lone AF (age<60 years, left atrial size <2.1 cm/m^2^ , no associated cardiopulmonary disease except mitral valve prolapse) - Patient or physician refuses - Age > 75 years (for patients enrolled before Nov 1988) | Jun 1987 to Nov 1989 |
| SPAF-2^24^ | AF on ECG (constant or intermittent) in the preceding 12 months | Same as reported for SPAF-1 | Sept 1985 to 1992 |
| SPAF-3^25^ | AF on ECG within 6 months  1 or more high risk features:   - Impaired left ventricle manifested by congestive heart failure within 100 days or fractional shortening ≤ 25% - Systolic blood pressure >160 mm Hg on one reading and >150 mm Hg on second reading or documented systolic blood pressure >160 mm Hg in preceding 3 months - Prior ischemic stroke, TIA, or systemic embolism more than 30 days prior to entry - Female and age over 75 years | Same as reported for SPAF-1 | May 1993 to Oct 1995 |
| Stroke Prevention in Non-rheumatic Atrial Fibrillation (SPINAF)^26^ | Male  No echocardiographic evidence of rheumatic heart disease  AF on 2 ECGs at least 4 weeks apart  Normal prothrombin time ratio | Intermittent AF  Definite indication for anticoagulant or antiplatelet  Chronic alcoholism, social or psychological condition unsuitable to anticoagulation  Co-existing medical disorder  Hemostasis disorder  Peptic ulcer disease within 2 years, esophageal varices, history of intracranial hemorrhage  History of gastrointestinal hemorrhage within 2 years  Planned surgery or invasive procedure  Lab abnormalities (Hct <32%, platelet count <100,000 /mm3, AST, ALT, or alkaline phosphatase > 2x ULN, guaiac-positive stool, > 5 RBCs/hpf in urine  Uncontrolled hypertension >180/105 mm Hg  Bacterial endocarditis  Atrial tumor  Anticoagulation within 6 months ago  Use of nonsteroidal anti-inflammatory  Uninterpretable echocardiogram  TIA within 5 years  Previous cerebral infarction  Hyperthyroidism  Planned cardioversion  Unstable angina | Jun 1987 to May 1993 |

Legend: AF – atrial fibrillation, ECG – electrocardiogram, SBP – systolic blood pressure, DBP – diastolic blood pressure, BP – blood pressure, TIA – transient, ischemic attack, NSAID – nonsteroidal anti-inflammatory drug, NYHA – New York Heart Association, ULN – upper limit of normal

### Appendix 2: Trial arms and enrollment

| **Trial** | **Mean observation time** | **Anticoagulant arm(s)** | **Antiplatelet arm** | **Combination** | **Placebo arm** |
| --- | --- | --- | --- | --- | --- |
| Atrial Fibrillation, Aspirin, and Anticoagulation Study 1 (AFASAK-1)^15^ | 1.3 yrs | Dose adjusted warfarin with goal INR 2.8-4.2 (N = 335) | Aspirin 75 mg/d  (N = 336) | n/a | N = 336 |
| AFASAK-2^16^ | 1.6 yrs | Fixed dose warfarin 1.25mg/d (N=167)*  Dose-adjusted warfarin with goal INR 2.0-3.0 (N=170) | Aspirin 300 mg/d (N=169) | Fixed-dose warfarin and aspirin 300 mg/d (N=171)† | n/a |
| Boston Area Anticoagulation Trial for Atrial Fibrillation (BAATAF)^17^ | 2.2 years | Dose-adjusted warfarin with goal INR 1.5-2.7 (N=212) ‡ | n/a | n/a | N=208 |
| Birmingham Atrial Fibrillation Treatment of the Aged Study (BAFTA)^18^ | 2.7 years | Dose-adjusted warfarin with goal INR 2.0-3.0 (N=488) | Aspirin 75 mg/d (N=485) | n/a | n/a |
| Canadian Atrial Fibrillation Anticoagulation (CAFA)^19^ | 1.3 years | Dose-adjusted warfarin with goal INR 2.0-3.0 (n=187) | n/a | n/a | N=191 |
| European Atrial Fibrillation Trial (EAFT)^20^  Group 1 (eligible for anticoagulants) § | 2.3 years | Dose-adjusted warfarin with goal INR 2.5-4.0 (N=225) | Aspirin 300 mg/d (N= 230) | n/a | N=214 |
| Primary Prevention of Arterial Thromboembolism in Atrial Fibrillation (PAATAF)^21^  Group 1 (eligible for anticoagulants) § | 3.0 years | Dose-adjusted warfarin with goal INR 2.5-3.5 (N=131)  Low-intensity coumarin with INR goal 1.1-1.6 (N=122) ‖ | Aspirin 150 mg/day (N=141) | n/a | n/a |
| National Study for Prevention of Embolism in Atrial Fibrillation (NASPEAF)^22^ ¶ | 2.4 years | Dose-adjusted acenocumarol with goal INR 2.0-3.0 (N=323) | Triflusal 600 mg/day (N=235)** | n/a | n/a |
| Stroke Prevention in Atrial Fibrillation 1 study (SPAF-1)^23^  Group 1 (eligible for anticoagulants) § †† | 1.2 years |  |  |  | N=211 |
| SPAF-2^24^ | 2.7 years | Dose-adjusted warfarin with goal INR 2.0-4.5 (N=555) | Aspirin 325 mg/d (N=545) | n/a | n/a |
| SPAF-3^25^ | 1.1 years | Dose adjusted warfarin with goal INR 2.0 to 3.0 (N=523) |  | Low, fixed dose warfarin (0.5 to 3.0 mg/d) to raise INR to between 1.2 and 1.5 and aspirin 325 mg/d (N=521) ‡‡ |  |
| Stroke Prevention in Non-rheumatic Atrial Fibrillation (SPINAF)^26^ | 1.7 years | Warfarin with goal INR 1.5 to 2.7‡ (N=265) |  |  | N=260 |

Legend

INR – international normalized ratio

* Trial reported a mean INR of 1.14, supporting categorization as control

† Trial reported a mean INR of 1.12, supporting categorization as control

‡ INR estimate prothrombin-time ratio

§ Group 2 were those deemed ineligible for anticoagulants and were thus excluded from the current analysis

‖ Trial reported a mean INR of 1.4 supporting categorization as control

¶ NASPEAF included patients with valvular atrial fibrillation, sample size reported here excludes those with valvular AF. Combination arms were excluded because the trial reported a median INR of 1.9 in the intermediate-risk group, and 2.1 in the high-risk group.

** Similar clinical efficacy to aspirin 300 mg/day

†† SPAF1 participants randomized to anticoagulation and aspirin were followed after the termination of SPAF1 and their results are reported in SPAF-2

‡‡ Mean INR in the fixed-dose group was 1.3 supporting categorization as control

### Appendix 3: Cohort flow diagram

Final cohort

(n= 7933)

Excluded due to missing data where the data element was otherwise collected in a trial (n=69)

- Missing height (n=41)
- Missing weight (n=27)
- Missing smoking status (n=6)
- Missing hypertension status (n=5)
- Missing diabetes status (n=4)
- Missing vascular disease status (n=3)
- Missing angina status (n=3)
- Missing heart failure status (n=1)
- Missing stroke history (n=1)
- Missing myocardial infarction history (n=1)
- Missing disenrollment date (n=1)

Randomized patients who met inclusion criteria (n= 8002)

### Appendix 4: Primary Trial Outcome Definition

| **Trial** | **Ischemic stroke** | **Systemic embolism** |
| --- | --- | --- |
| Atrial Fibrillation, Aspirin, and Anticoagulation Study 1 (AFASAK-1)^15^ | Clinical signs or a medically confirmed history of acute onset of a neurological deficit of presumed vascular origin lasting >24hrs. CT when possible; death certificates and autopsy reports for fatal strokes. | Embolism to viscera or extremities based on symptoms relevant diagnostics test, surgery, or autopsy |
| AFASAK-2^16^ | Acute focal neurological deficit of presumed vascular origin >24 hours; no blood on CT/MRI | Embolism to kidney, spleen, gut, lung, or extremities by angiography, surgery, or autopsy |
| Boston Area Anticoagulation Trial for Atrial Fibrillation (BAATAF)^17^ | Focal neurologic deficit >24 hours without blood on CT | Assessed as an outcome but not defined; no events reported |
| Birmingham Atrial Fibrillation Treatment of the Aged Study (BAFTA)^18^ | Focal neurological deficit >24 hours without blood on CT | Embolism to kidney, spleen, gut, or extremities by angiography, scintigraphy, surgery, or autopsy |
| Canadian Atrial Fibrillation Anticoagulation (CAFA)^19^ | Focal neurologic deficit >24 hours with new, appropriate finding on CT, excluding lacunar events | Embolism to gut, kidney, or extremities by angiography or surgery |
| European Atrial Fibrillation Trial (EAFT)^20^ | Focal neurologic deficit >24 hours without blood on CT | Sudden vascular insufficiency of limbs or internal organs associated with evidence of arterial occlusion, in the absence of previous obstructive disease |
| Primary Prevention of Arterial Thromboembolism in Atrial Fibrillation (PAATAF)^21^ | Focal neurologic deficit >24 hours without blood on CT | Acute vascular occlusion resulting in recovery, permanent sequelae, or death |
| National Study for Prevention of Embolism in Atrial Fibrillation (NASPEAF)^22^ | Focal neurologic deficit >24 hours without blood on neuroimaging | Abrupt vascular insufficiency without previous symptoms |
| Stroke Prevention in Atrial Fibrillation 1 study (SPAF-1)^23^ | Acute focal neurological deficit in the distribution of a single brain artery lasting >24 hours; no blood on CT/MRI | Abrupt vascular insufficiency associated with clinical or radiologic evidence of arterial occlusion in the absence of other likely mechanisms (e.g., atherosclerosis, instrumentation). |
| SPAF-2^24^ | Focal neurological deficit of sudden onset had to persist beyond 24 hours; ischemic cause was confirmed by neuroimaging or autopsy | Abrupt vascular insufficiency related to arterial occlusion without previous obstructive disease. |
| SPAF-3^25^ | Focal neurological symptoms or signs of sudden onset persisting for more than 24 h; the absence of primary hemorrhage was confirmed by neuroimaging or autopsy | Abrupt vascular insufficiency related to arterial occlusion without previous obstructive disease. |
| Stroke Prevention in Non-rheumatic Atrial Fibrillation (SPINAF)^26^ | Focal neurologic deficit >12 hours without blood or tumor on CT | Not considered a primary or secondary outcome |

### Appendix 5 Estimation of life expectancy

#### 5.1 Background

We estimated the life expectancy for each participant using the life table method.^47^ We started with the Centers for Disease Control and Prevention life tables. We selected the sex- and enrollment year-specific life table for each participant. We modified the expected annual mortality based on the excess mortality risk associated with their baseline comorbidities for each year following trial enrollment. The following equation estimated modified mortality risk:

$q_{xi}=1-e^{-1[\left( \sum_{1}^{n} n_{i}*z*q_{x} \right) + q_{x}]}$ (1)

Where $q_{xi}$ is the estimated 1-year mortality risk for the $i^{th}$patient at a given age on or after trial enrollment. $z$ represented the excess hazard of a patient comorbidity (indicator variable $n_{i}$) and $q_{x}$ is the CDC life table mortality risk based on the patient’s age, sex, and trial enrollment year. Values of $z$ are presented in Appendix Table 5.2. We chose specific comorbidities to adjust life expectancy based on our prior studies, baseline characteristics present in the AFI database, and conditions for which published epidemiological studies on excess mortality risk exist. We used this approach in prior analyses.^37^

#### 5.2 Table: Excess mortality risk for comorbidities adjusted for in this analysis

| **Condition** | **Subset** | **Excess mortality risk**  ($z$ in equation 1) |
| --- | --- | --- |
| Diabetes (Tancredi et al. NEJM 2015, Table 2) | < 55 years | 1.32 |
|  | Age >= 55, < 65 years | 0.72 |
|  | Age >= 65,< 75 years | 0.42 |
|  | Age >= 75 years | 0.11 |
| Heart Failure (Roger et al. JAMA 2004 Table 3 5-yr mortality standardized against age, sex, year-specific population mortality) | Male 1979 to 1984 | 0.79 |
|  | Female 1979 to 1984 | 1.52 |
|  | Male 1985 to 1990 | 1.10 |
|  | Female 1985 to 1990 | 2.11 |
|  | Male 1991 to 1995 | 1.27 |
|  | Female 1991 to 1995 | 2.38 |
|  | Male after 1996 | 1.10 |
|  | Female after 1996 | 2.05 |
| Stroke (Brønnum al. Stroke 2001 Table 2) | Male >= 70 years | 0.92 |
|  | Male < 70 years | 2.01 |
|  | Female >= 70 years | 1.05 |
|  | Male < 70 years | 2.52 |
| Body Mass Index (Flegal et al. 2005 JAMA; Table 2) | BMI < 18.5 and (Age >=60, < 70 years) | 1.30 |
|  | BMI 18.5 and (Age >= 70 years) | 0.69 |
|  | BMI > 35 and (Age < 60 years) | 0.83 |
|  | BMI > 35 and (Age >= 60, < 70 years) | 0.63 |
| Tobacco use (Carter et al. NEJM 2015; supplemental appendix C) | Current use | 1.80 |
|  | Former use | 0.50 |
| Acute myocardial infarction (Norgaard et al. Diabetologia 2010; Table 2) | Male before 2001 | 0.40 |
|  | Female before 2001 | 0.87 |
|  | Male 2001 or after | 0.465 |
|  | Female 2001 or after | 0.91 |

#### 5.3 Worked example

Consider an 80-year-old man enrolled in a trial in 1990 with a history of an acute myocardial infarction and diabetes. The baseline 1-year mortality risk for an 80-year-old male in 1990 ($q_{x}$) is 0.08542 from the CDC life table. Based on equation (1) above, this patient’s individualized mortality risk ($q_{xi})$ would be calculated as:

$$q_{xi}=1-e^{-1[ (0.40*0.08542)+(0.11*0.08542)+(0.08542)]}$$

$$q_{xi}=0.12101$$

We perform the same calculation for each year for each patient following trial enrollment. Updated mortality rates are then used to calculate the patient’s estimated life expectancy at trial enrollment using the life table method.

#### 5.4 Performance of life expectancy compared to age

In the final study cohort, life expectancy and age are inversely correlated (Pearson correlation coefficient of -0.77). To determine their relative performance, we fit two separate Cox models with time to death as the outcome. The first included age as the sole predictor and the other with life expectancy as the sole predictor. We found that life expectancy has a better discriminatory capacity when predicting death at 3 years in the AFI cohort (Harrell’s c-statistic 0.68 vs. 0.61).

### Appendix 6: Estimation of absolute risk reduction over multiple years using the AHA/ACC/HRS method

When extrapolating the annual risk reduction over multiple year, one must account for the declining population at risk. To do this, the following formula is usually employed:

$$ARR in year Y=1-\exp\left( -annual ARR*Y \right)$$

For our use case, we substitute $annual stroke rate*0.64$ for $annual ARR$. Annual off-treatment stroke rate is determined from the CHA_2_DS_2_-VASc score and 0.64 is the efficacy of anticoagulants to prevent stroke.

As a result we used the following formula

$$ARR in year Y=1-\exp\left( -annual stroke rate*0.64*Y \right).$$

This is the same approach and formula used in clinical decision aids (e.g., Zeballos-Palacios et al. Mayo Clin Proc. 2019;94(4):686-696)

### Appendix 7: Table 1 Baseline characteristic after 20 chained imputations

| **Characteristic** |  |
| --- | --- |
| Age, years (median [IQR]) | 73 [67, 78] |
| Life expectancy, years (median [IQR]) | 8 [6, 12] |
| Sex |  |
| Male | 64% |
| Female | 36% |
| Diabetes |  |
| No | 85% |
| Yes | 15% |
| Hypertension |  |
| No | 51% |
| Yes | 49% |
| Congestive heart failure |  |
| No | 70% |
| Yes | 30% |
| Prior stroke |  |
| No | 81% |
| Yes | 19% |
| Angina |  |
| No | 83% |
| Yes | 17% |
| Prior myocardial infarction |  |
| No | 88% |
| Yes | 12% |
| Peripheral vascular disease |  |
| No | 92% |
| Yes | 8% |
| Body mass index kg/m^2^ (median [IQR]) | 26 [24, 29] |
| Smoking status |  |
| Never smoker | 49% |
| Former smoker | 36% |
| Current smoker | 16% |
| CHA_2_DS_2_-VASc score (median [IQR]) | 3 [2, 4] |
| Trial |  |
| AFASAK1 | 13% |
| BAATAF | 5% |
| CAFA | 5% |
| SPAF1 | 3% |
| SPINAF | 7% |
| EAFT | 8% |
| PATAF | 5% |
| SPAF2 | 14% |
| AFASAK2 | 9% |
| SPAF3 | 13% |
| NASPEAF | 7% |
| BAFTA | 12% |
| Randomized study arm assignment |  |
| Warfarin | 43% |
| Control | 57% |

### Appendix 8: Fine-Gray model parameters for Competing Risk Model

Time to stroke or systemic embolism is the primary outcome and death is treated as a competing event. All CHA_2_DS_2_-VASc predictors are also included.

| **Parameter** | **HR** | **LL** | **UL** |
| --- | --- | --- | --- |
| Age (65-74 years, reference <65 years) | 1.17 | 0.95 | 1.45 |
| Age (75+ years, reference <65 years) | 1.59 | 1.17 | 2.15 |
| Women | 1.15 | 0.97 | 1.35 |
| History of congestive heart failure | 1.12 | 0.89 | 1.42 |
| History of hypertension | 1.40 | 1.20 | 1.65 |
| History of diabetes | 1.40 | 1.21 | 1.62 |
| History of prior stroke or TIA | 3.13 | 2.45 | 4.00 |
| History of peripheral vascular disease | 1.13 | 0.91 | 1.40 |
| Randomization to anticoagulant arm | 0.39 | 0.31 | 0.49 |

### Appendix 9: Comparing ARR estimated by CHA_2_DS_2_-VASc with an internally re-calibrated CHA_2_DS_2_-VASc ARR

#### 9.1 Background

To calculate absolute risk reduction in the main analysis, we used the procedure detailed in the “2014 AHA/ACC/HRS Guideline for the Management of Patients With Atrial Fibrillation.” This procedure is also used in decision aids developed by the ACC and Mayo Clinic. In brief, this method uses annual stroke rates corresponding to each CHA_2_DS_2_-VASc score from Friberg et al. (European Heart Journal (2012) 33, 1500–1510) and then apply a 0.64 relative risk quoted in the 2014 AHA/ACC/HRS guidelines. Together these two studies are used to estimate the absolute risk reduction attributable to therapy in guidelines and decision tools.

The main analysis compared the guideline estimate of absolute risk reduction to a Fine-Gray model that treats death as a competing event, which is estimated in the AFI database. Since the guideline estimates were developed in external cohorts, they may be miscalibrated resulting in the over-estimate of treatment benefit observed in the main analysis.

In this sensitivity analysis, we determined if the CHA_2_DS_2_-VASc would perform better if it were re-estimated in the AFI database. We estimate CHA_2_DS_2_-VASc specific stroke rates by dividing the number of events by person time. We then apply the hazard ratio of treatment from this analysis to determine the absolute risk reduction. This approach mirrors the current guidelines approach but is recalibrated.

The sensitivity analysis shows that recalibrating the absolute risk reduction by CHA_2_DS_2_-VASc score in the AFI database does not meaningfully change the primary conclusion. In fact, the sensitivity analysis showed that recalibration resulted in greater differences in estimates when compared to the Competing Risk Model.

#### 9.2 Comparison of stroke/systemic embolism rates and absolute risk reduction with anticoagulants per 100 person-years re-estimated in the AFI database

| CHA2DS2-VASc score* | **Guidelines reported expected event rates**  **(used in main analysis)** | | **Event Rates from AFI** | |
| --- | --- | --- | --- | --- |
|  | Ischemic Stroke/systemic embolism rate,  off-treatment (Friberg 2012)** | Ischemic Stroke/systemic embolism rate,  on-treatment by guidelines | Ischemic Stroke/systemic embolism rate recalibrated in AFI data, off-treatment  (95% CI) | Ischemic Stroke/systemic embolism rate,  on-treatment re-estimated in AFI data |
| 0 | 0.2 | 0.07 | 0.44 (0.07 to 1.46) | 0.17 |
| 1 | 0.6 | 0.22 | 1.48 (0.97 to 2.16) | 0.58 |
| 2 | 2.5 | 1.60 | 3.40 (2.67 to 4.27) | 1.32 |
| 3 | 3.7 | 1.33 | 4.43 (3.66 to 5.36) | 1.73 |
| 4 | 5.5 | 1.98 | 6.12 (5.08 to 7.34) | 2.39 |
| 5 | 8.4 | 3.02 | 9.30 (7.52 to 11.53) | 3.63 |
| 6 | 11.4 | 4.10 | 10.9 (8.1 to 14.6) | 4.27 |
| 7 | 13.1 | 4.72 | 12.8 (6.2 to 23.5) | 5.01 |
| 8 | 12.6 | 4.54 | 23.2 (7.4 to 56.0) | 9.04 |

Legend

*no patients in the AFI database had a CHA_2_DS_2_-VASc score of 9

** Friberg 2012 did not present 95% confidence intervals, or the data needed to calculate confidence intervals

#### 9.3 Table Estimated ARR by CHA_2_DS_2_-VASc model compared with Competing Risk Model and recalibrated CHA_2_DS_2_-VASc (replication of Figure 1)

| **Years after randomization** | **ARR by Competing Risk Model**  **(Main analysis Fig 1)** | **ARR by CHA2DS2-VASc**  **(Main analysis Fig 1)** | **ARR by recalibrated CHA2DS2-VASc (Sensitivity analysis)** |
| --- | --- | --- | --- |
| 1 | 2.4% [IQR 1.8% to 3.6%] | 2.3% [IQR 1.6% to 3.5%] | 2.7% [IQR 2.1% to 3.7%] |
| 2 | 4.0% [IQR 2.9% to 5.8%] | 4.6% [IQR 3.1% to 6.8%] | 5.3% [IQR 4.1% to 7.2%] |
| 3 | 5.2% [IQR 3.8% to 7.4%] | 6.9% [IQR 4.7% to 10.0%] | 7.8% [IQR 6.0% to 10.6%] |
| 4 | 6.2% [IQR 4.5% to 8.7%] | 9.0% [IQR 6.2% to 13.1%] | 10.2% [IQR 8.0% to 13.9%] |
| 5 | 6.3% [IQR 4.6% to 8.9%] | 11.2% [IQR 7.7% to 16.1%] | 12.6% [IQR 9.9% to 17.0%] |

### Appendix 10: Estimated ARR by CHA_2_DS_2_-VASc model compared to Competing Risk Model

#### 10.1 Figure 1 in tabular format

| Time horizon (years) | CHA_2_DS_2_-VASc model ARR,  median (IQR) | Competing Risk Model ARR, median (IRQ) |
| --- | --- | --- |
| 1 | 2.3% (IQR 1.6%, 3.5%) | 2.4% (IQR 1.8%, 3.6%) |
| 2 | 4.6% (IQR 3.1%, 6.8%) | 4.0% (IQR 2.9%, 5.8%) |
| 3 | 6.9% (IQR 4.7%, 10.0%) | 5.2% (IQR 3.8%, 7.4%) |
| 4 | 9.0% (IQR 6.2%, 13.1%) | 6.2% (IQR 4.5%, 8.7%) |
| 5 | 11.2% (IQR 7.7%, 16.1%) | 6.3% (IQR 4.6%, 8.9%) |

#### 10.2 Estimated cumulative incidence of stroke or systemic embolism by model and treatment status

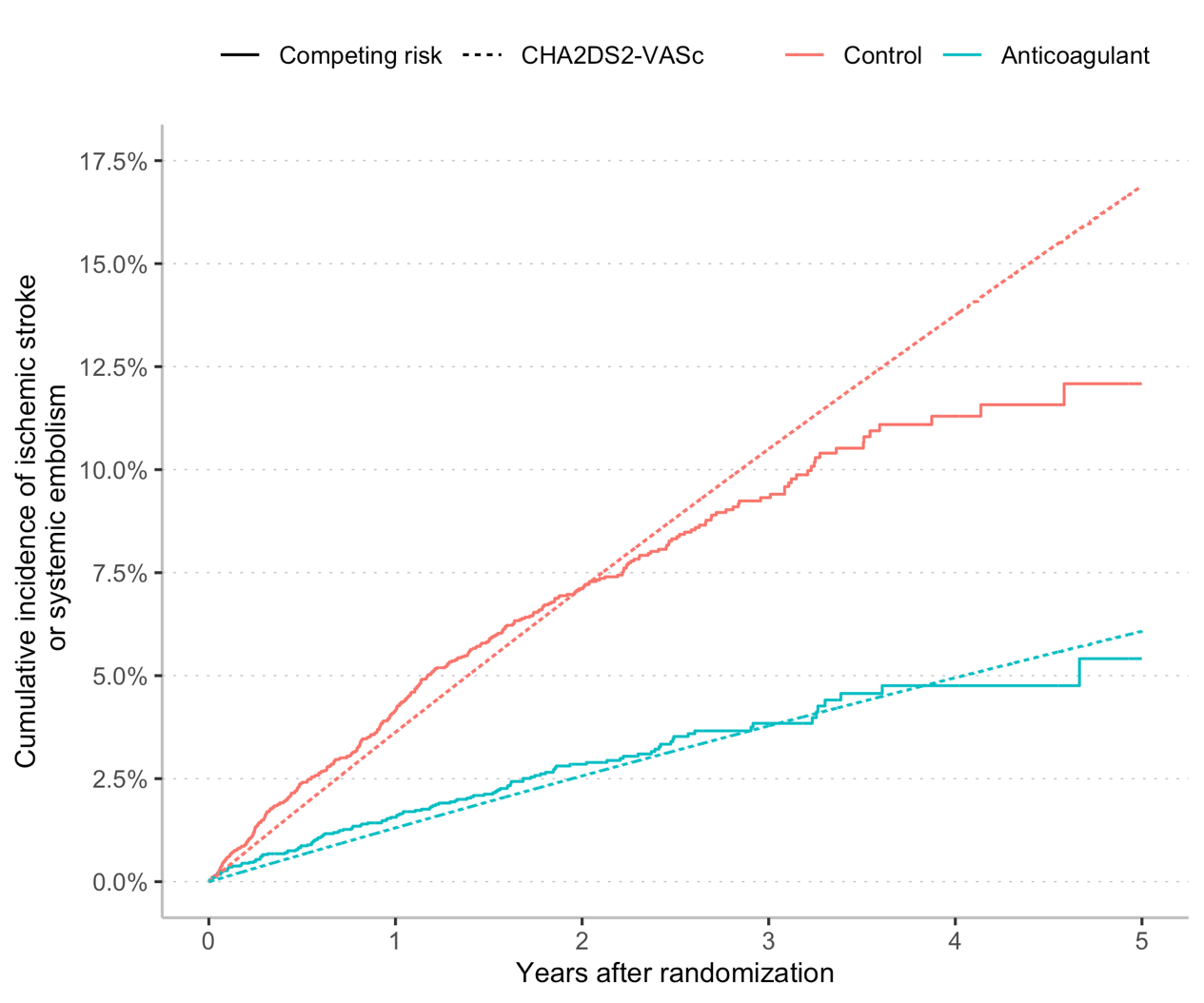

### Appendix 11: Misestimation of ARR by CHA_2_DS_2_-VASc score at 3 years (Figure 2 in tabular format)

| **Decile of life expectancy (range)** | **Absolute misestimation of CHA_2_DS_2_-VASc estimate at 3 years, mean (95% CI)** | **Relative misestimation of CHA_2_DS_2_-VASc estimate at 3 years (95% CI)** |
| --- | --- | --- |
| 1 (16 to 47 yrs.) | -1.3% (-1.3% to -1.2%) | -44% (-46% to -41%) |
| 2 (13 to 16 yrs.) | -1.0% (-1.1% to -0.9%) | -27% (-30% to -25%) |
| 3 (11 to 13 yrs.) | -0.3% ( -0.5% to -0.1%) | -7% (-10% to -4%) |
| 4 (9 to 11 yrs.) | 0.1% ( -0.1% to 0.3%) | 5% (2% to 8%) |
| 5 (8 to 9 yrs.) | 0.3% (0.1% to 0.5%) | 9% (6% to 12%) |
| 6 (7 to 8 yrs.) | 0.5% (0.2% to 0.7%) | 17% (15% to 20%) |
| 7 (6 to 7 yrs.) | 0.8% (0.6% to 1.0%) | 23% (20% to 25%) |
| 8 (5 to 6 yrs.) | 1.8% (1.5% to 2.0%) | 37% (34% to 41%) |
| 9 (4 to 5 yrs.) | 3.0% (2.7% to 3.2%) | 58% (54% to 62%) |
| 10 (1 to 4 yrs.) | 4.7% (4.5% to 5.0%) | 78% (74% to 82%) |
